# Pulsatility analysis of the circle of Willis

**DOI:** 10.1101/2024.02.13.24302767

**Authors:** Henning U. Voss, Qolamreza R. Razlighi

## Abstract

**Purpose:** To evaluate the phenomenological significance of cerebral blood pulsatility imaging in aging research.

**Methods:** N = 38 subjects aged from 20 to 72 years of age (24 females) were imaged with ultrafast MRI with a sampling rate of 100 ms and simultaneous acquisition of pulse oximetry data. Of these, 28 subjects had acceptable MRI and pulse data, with 16 subjects between 20 and 28 years of age, and 12 subjects between 61 and 72 years of age. Pulse amplitude in the circle of Willis was assessed with the recently developed method of analytic phase projection to extract blood volume waveforms with an effective sampling rate of about 0.3 ms.

**Results:** Arteries in the circle of Willis showed pulsatility in the MRI for both the young and old age groups. Pulse amplitude in the circle of Willis significantly increased with age (p = 0.01) but was independent of gender, heart rate, and head motion during MRI.

**Discussion and conclusion:** Increased pulse wave amplitude in the circle of Willis in the elderly suggests a phenomenological significance of cerebral blood pulsatility imaging in aging research. The physiologic origin of increased pulse amplitude (increased pulse pressure vs. change in arterial morphology vs. re-shaping of pulse waveforms caused by the heart) requires further investigation.

## Introduction

Cerebrovascular pulsatility has gained significant importance as a research topic, primarily due to its potential impact on aging-related brain health. With advancing age, and in certain conditions such as chronic hypertension, the physical properties of the brain vasculature, and subsequently the vascular network, including its viscoelasticity, undergo changes^1-5^. These changes can lead to a reduction of the absorption of blood pressure pulsations, an increased impedance, and the potential for damage, including hemorrhagic stroke and microbleeds. Notably, small-scale changes that may not be evident in conventional imaging have attracted attention as potential contributors to conditions like dementia^6-8^. Furthermore, there is a growing body of evidence linking cerebrovascular health to various aspects of cognitive function and neurodegenerative diseases. For instance, aortic stiffness has been associated with cerebral small-vessel disease in hypertensive patients^9^, cognitive decline correlates with the pulse wave velocity^10^, and neurodegenerative diseases such as Alzheimer’s disease are believed to be connected to cerebrovascular dysregulation^11-15^. Additionally, central artery stiffness has been reported to impact the perfusion of deep subcortical matter^16^, central arterial aging appears to be linked to an increased volume of white matter hyperintensities^17^, and arterial stiffness is associated with beta-amyloid deposition in the elderly^18^. In a more indirect manner, the pressure gradient induced by arterial pulsatility could potentially serve as a driving force in the brain’s paravascular waste clearance system,^19, 20^ which plays a crucial role in brain health by facilitating the removal of metabolic waste products..

Therefore, it would be advantageous to directly image vascular changes in the brain. However, there are only limited *in vivo* options for imaging vascular dynamics in the human brain. Arterial spin labelling is probably the most advanced current method with the highest spatial resolution^21, 22^. It provides quantitative blood flow values in the capillary bed, assuming steady flow^23^. In contrast, the important parameter of arterial compliance depends on the pulsatile component of the flow. It can be imaged for larger vessels locally with transcranial Doppler ultrasound^24, 25^. Pulsatile flow components can also be imaged over the whole brain with 4D phase contrast angiography^26, 27^. However, there has been recent progress in imaging pulsatility^28-37^ in the brain with fast echo-planar imaging (EPI). It is important to note that in the main cerebral arteries, pulse wave velocity is typically higher in magnitude (up to about 12 m/s)^38^ than blood flow velocity (about 30 to 100 cm/s; varies widely with age, cardiac phase, blood pressure, type of artery, and other factors^39, 40^.

In this contribution, we utilize the method of MRI hypersampling by analytic phase projection^41^, which has been proven to reveal pulse waveforms in the main cerebral arteries. Importantly, this method can be seamlessly integrated into clinical MRI scans without the requirement for non-standard equipment or pulse sequences.

The region under study is the circle of Willis (COW), chosen for both pragmatic and scientific reasons. Pragmatically, the COW is easy to define in MRI, and for the sake of ultrafast imaging, it provides a suitable region with little inter-subject variability due to reproducible slice prescription. From a scientific standpoint, the COW holds a significant role in pulsatility, as it has been hypothesized to function as a pressure absorber mechanism that helps prevent damage to the cranial microvasculature and the blood-brain barrier^42^. However, it is worth noting that everything in the skull pulsates in synchrony with the heartbeat^42-46^, including cerebrospinal fluid, the cortical surface, and veins. The methods used in this study could potentially be useful for investigating pulsatility and pulse waves in other areas of the brain as well.

The research presented here is motivated by our enthusiasm for the exciting quest to better understand the aging human brain via biomedical imaging^47-61^.

## Materials and Methods

### Subjects

All participants were recruited using random market mailing approach within 50-mile radius of Columbia University Irving Medical Center and were compensated for their time spent taking part in this study. All subjects gave their informed written consent prior to the scanning sessions. The experimental design of our study and the recruitment process were approved by Columbia University institutional review board. Thirty-eight young and older but otherwise healthy participants (with cognitive scores within 2 standard deviations of the age-matched norm) between 20 to 72 years of age (24 female) were recruited. The MRI and pulse data were inspected for quality, and 28 subjects had acceptable MRI and pulse data. Specifically, the ten scans that could not be used for further analysis consisted of two scans that had an incorrect acquisition plane orientation, four scans that showed extreme head motion, and four scans that did not have usable pulse data. Of the remaining scans used for further analysis, 16 subjects were between 20 and 28 years of age (12 female) and 12 subjects between 61 and 72 years of age (5 female).

### Imaging

Subjects were imaged using a 3 Tesla Siemens Magnetom Prisma scanner equipped with an 80 mT/m gradient system with a T_2_^*^-weighted ultrafast echo-planar imaging (EPI) sequence (repetition time =100 ms, echo time = 35 ms, field-of-view of 22 cm, matrix size of 100 × 100, and slice thickness of 3 mm) with simultaneous and synchronized acquisition of pulse oximetry data from the index finger of the left hand. One slice passing through the COW was imaged with 3000 repetitions totaling 5 min scan time.

### Pulse waveform analysis

#### Preprocessing of raw MRI data

The first fMRI image of each subject was used as an anchor to which all subsequent 2999 images were aligned to by an intensity-based rigid image registration algorithm (MATLAB’s imregister function. The EPI time series were detrended with a zero-lag highpass filter with a cutoff at 20 s, corresponding to 200 data points.

#### Preprocessing of pulse oximetry data

The pulse oximetry signals were filtered with a zero-lag lowpass filter with a cutoff at 1.5 Hz to enhance pseudo-periodicity^62-64^ of the pulse signal, and to remove noise.

#### Estimation of pulse wave form and amplitude

For each subject, MRI pulse waveforms were extracted from the EPI-MRI data by analytic phase projection (APP)^41, 65, 66^. APP is a generalized form of retrospective gating. It overcomes the arbitrary limitation that the phase within an inter-beat interval is assumed to increase linearly: In APP, the phase of the pulse oximetry signal is estimated from the analytic phase of the pulse oximetry signal via a Hilbert transform. Thus, it can be nonlinear, leading to a more accurate gating for variable inter-beat intervals^67^. Then, each MRI data point is mapped to its corresponding phase, leading to a reordering of MRI data points in time, analogously to retrospective gating. Both retrospective gating and APP lead to an effective sampling interval that is much smaller than the TR of the MRI acquisition. For example, if the average inter-beat interval has period T and N MRI images are sampled, the effective hypersampling time is T/N. In the present study, this amounts to a hypersampling time of approximately 0.3 ms. The pulse oximetry signals were then shifted in time to match the MRI acquisition sample times. The amount of shift was estimated by searching for the maximum average pulse signal amplitude in the hypersampled MRI data. The average pulse signal amplitude was averaged from the amplitudes in each voxel in the brain, by using a hand-drawn mask that excluded ventricles and skull areas. Finally, pulse waveforms were estimated by smoothing the hypersampled MRI signal with a zero-lag filter, retaining six cycles per cardiac cycle, and subsequent amplitude normalization.

#### Motion assessment

For each subject, head motion was quantified by the standard deviation of the rotation angle of the rigid transformation used for motion correction, using MATLAB’s imregtform function. In order to exclude the possibility that the amount of head motion during MRI scanning biased pulsatility measures, this parameter was included as a covariate in the statistical analysis.

### Statistics

Two circular regions of interest (ROIs) were defined for each fMRI data set (Figure 1): 1. An ROI centered at the COW, and 2. an ROI centered in a brain area nearby without noticeable pulsatility, used as baseline. The MRI pulse amplitude was computed for these ROIs as the average of the differences between the maximum and minimum waveform value. In the COW, this amplitude was then divided by the median amplitude from the baseline ROI. To evaluate the age dependence of pulsatility, a multiple regression analysis was performed with MRI pulse amplitudes in the COW as dependent variable and age, heart rate, motion, and gender as independent variables. Gender was used as a categorical variable. In addition, another multiple regression analysis was used to test for the possibility that the baseline ROI depends on any of the three independent variables.

**Figure 1:**
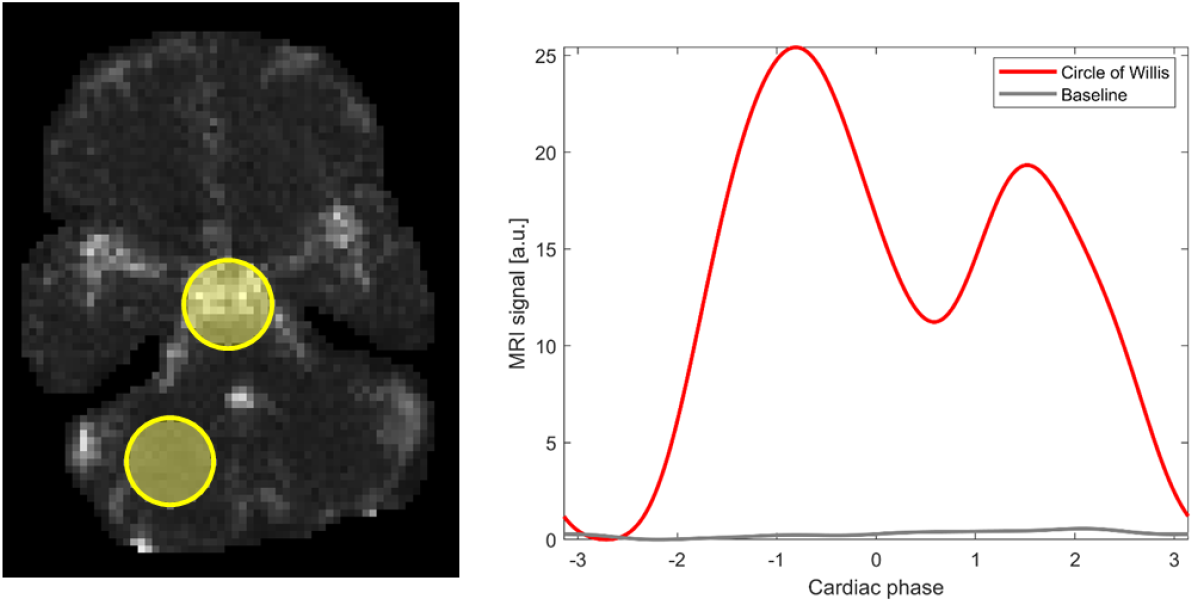
MRI pulse waves. Left: The square root of MRI pulse wave amplitudes over the brain slice acquired with ultrafast MRI, for a single subject. The region of interest for the analysis of the circle of Willis and the baseline region without dominant pulsatility are highlighted. Middle: The maximum-amplitude MRI pulse wave in the circle of Willis shows the characteristic systolic upstroke starting at around phase value -2.5 rad and the subsequent diastolic parts. In addition, the maximum-amplitude baseline signal is shown in gray.

## Results

Pulse amplitude maps could reliably be obtained from all 28 subjects and showed pulsatility in the circle of Willis, parts of the middle cerebral arteries, arteries in the scalp, and other regions. An example pulse amplitude map is shown in Fig. 1 for a single subject. Estimates of pulse waveforms often showed characteristic features such as the systolic upstroke and dicrotic notch.

A multiple regression analysis revealed that pulse amplitude in the COW increased with age (p = 0.01) but was not significantly dependent on heart rate, gender, or head motion during MRI. In the baseline region, pulse amplitude did not depend on age, heart rate, motion, or gender. The statistics is summarized in Table 1, and the increase of pulse amplitude with age in the COW is shown in Figure 2.

**Table 1:**
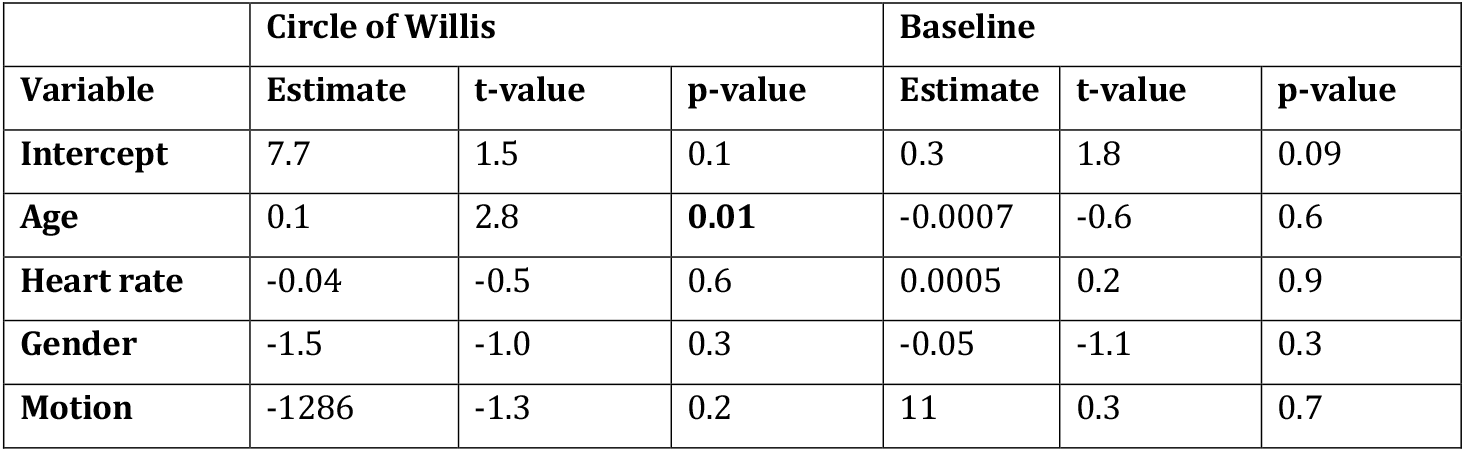
Multiple linear regression analysis.

**Figure 2:**
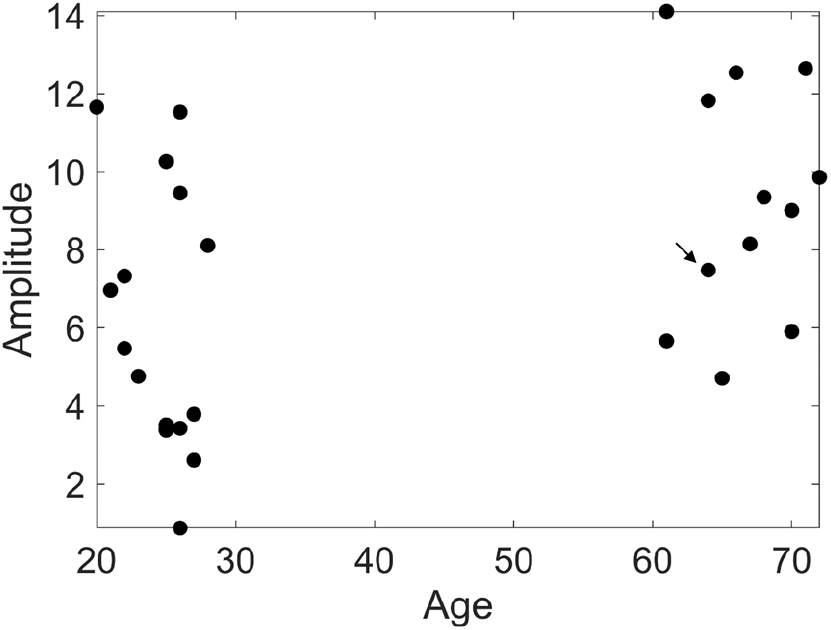
Increase of mean pulse amplitudes with age in the circle of Willis. The observed increase with age is significant with a p-value of 0.013 (post-hoc non-parametric Wilcoxon test). The amplitudes used here were normalized by the median baseline amplitude for each subject. The subject of Fig. 1 is marked with an arrow.

## Discussion

Our interpretation of age-related increase of fMRI derived pulse wave amplitude in the circle of Willis is as follows. In general, arterial blood flow consists of two components: The steady and the pulsatile flow^68-70^. Outside the capillary bed^71^, the former is mainly affected by peripheral vascular resistance, while the latter is mainly affected by arterial stiffness. After the age of 60, increases in blood pressure are primarily attributable to a rapid increase of pulse pressure (the difference between systolic and diastolic pressure), driven by the increase in systolic pressure^72^. We suggest that increased pulse pressure in cerebral arteries causes increased pulse amplitude in the COW, because the pulsatile blood flow experiences a relatively greater increase than the steady flow. An alternative explanation for larger pulse wave amplitudes could be that the arterial compliance, the ratio of blood volume and blood pressure, increases with age in the circle of Willis. However, due to the lack of information about intracranial blood pressure, and the fact that this explanation would contradict the general trend of an increase of arterial stiffness with age, this seems to be less likely.

It is an open question whether the increase in waveform amplitude is due to a change of the wave form before it enters the brain, due to a change inside the cerebral arteries, or due to an altered wave reflection at the arteriole/capillary level. A more thorough investigation would have to involve the measurement of wave forms at the aortic or carotid level^73, 74^, and MRI of larger brain regions. It is neither clear from the present data if an actual re-shaping of the waves or an amplification, or a combined effect thereof, takes place^75-80^. For a detailed analysis of wave forms, one would have to acquire data with higher spatial resolution that would allow for probing specific vessels rather than the *average* pulse amplitude in the circle of Willis. With faster imaging methods and higher field MRI^81-85^, studies like these are now within reach and subject of further research.

## Conclusion

We have evaluated the significance of cerebral blood pulsatility imaging in the circle of Willis and investigated its changes with age. We showed that the arteries of the circle of Willis exhibit MRI-pulsatility for both the young and old age groups, and that pulse amplitude significantly increased with age. This suggests a phenomenological significance of cerebral blood pulsatility imaging in aging research. However, the physiologic origin of increased pulse amplitude requires further investigation.

## Data Availability

Data produced in the present study are available to scientific researchers upon reasonable request to the authors

## Acknowledgments

The authors acknowledge help from David Parker and Amirreza Sedaghat.

## Author contributions

H. U. Voss: Data curation; Formal analysis; Funding acquisition; Investigation; Methodology; Software; Writing - original draft.

O. R. Razlighi Conceptualization; Data curation; Investigation; Project administration; Resources; Supervision; Writing - review & editing

## Disclosure/Conflict of Interest

The corresponding author is the inventor of patent WO 2019/199960A1, awarded to Cornell University, that describes the method of hypersampling used in this work.

## Notes

### Funding Statement

This study did not receive any funding

### Author Declarations

The Institutional Review Board of Columbia University, New York, gave ethical approval for this work

